# Does screening for *Neisseria gonorrhoeae* and *Chlamydia trachomatis* affect the incidence of these infections in men who have sex with men taking HIV pre-exposure prophylaxis (PrEP)Results from a randomized, multicentre controlled trial (the Gonoscreen study)

**DOI:** 10.1101/2023.08.14.23294056

**Authors:** Thibaut Vanbaelen, Achilleas Tsoumanis, Eric Florence, Christophe Van Dijck, Diana Huis in ‘t Veld, Anne-Sophie Sauvage, Natacha Herssens, Irith De Baetselier, Anke Rotsaert, Veronique Verhoeven, Sophie Henrard, Yven Van Herrewege, Dorien Van den Bossche, Jean-Christophe Goffard, Elizaveta Padalko, Thijs Reyniers, Bea Vuylsteke, Marie-Pierre Hayette, Agnes Libois, Chris Kenyon

## Abstract

**Background:** Guidelines recommend three-site (urine, anal, pharynx) three-monthly (3X3 screening) screening for Neisseria gonorrhoeae (NG) and Chlamydia trachomatis (CT) in men who have sex with men (MSM) taking HIV pre-exposure prophylaxis (PrEP). We present the first randomized controlled trial to compare the effect of screening versus non-screening for NG/CT on the incidence of these infections in MSM taking PrEP.

**Methods:** A multicenter, randomized, controlled trial of 3X3 screening for NG/CT versus non-screening was conducted among MSM taking PrEP in five HIV reference centers in Belgium. Participants attended the PrEP clinics quarterly for 12 months. NG/CT was tested at each visit in both arms, but results were not provided to the non-screening arm. The primary outcome was the incidence rate (IR) of NG/CT infections in each arm, assessed in the per-protocol population. Non-inferiority of the non-screening arm was proven if the upper limit of the 95% confidence interval of the IR ratio (IRR) was lower than 1.25. The trial protocol was registered at clinicaltrials.gov (NCT04269434).

**Findings:** Between September 2020 and June 2021, 508 subjects were randomized to the 3X3 screening arm and 506 to the non-screening arm. The overall IR of NG/CT was 0.155 cases/100 person-days (95%CI 0.128-0.186) in the 3×3 screening arm and 0.205 (95%CI 0.171-0.246) in the non-screening arm. The IR was significantly higher in the non-screening arm (IRR 1.318, 95%CI 1.068-1.627). Participants in the non-screening arm had a higher incidence of CT infections and symptomatic CT infections. There were no significant differences in NG infections. Participants in the non-screening arm consumed significantly less antimicrobials. No serious adverse events were reported.

**Interpretation:** We failed to show that non-screening for NG/CT is non-inferior to 3-site 3-monthly screening in MSM taking PrEP in Belgium. However, screening was associated with higher antibiotic consumption and had no effect on the incidence of NG. Therefore, our findings do not provide strong support for screening for NG/CT in this population.

**Funding:** Belgian Healthcare Knowledge Center (KCE - INV18-1133)

## Research in context

### Evidence before this study

We searched PubMed until April 06, 2023 for reports of randomized, controlled, clinical trials reporting the effect of screening for *Neisseria gonorrhoeae* or *Chlamydia trachomatis* on the prevalence or incidence of these infections. We used the search terms “chlamydia” OR “gonorrh*” and “screening” OR “testing” and “trial”. We found no reports of such trials for *Neisseria gonorrhoeae*. We found two randomized controlled trials assessing the effect of population-level screening for *Chlamydia trachomatis*. A randomized, step-wedge, controlled trial explored the effect of yearly screening for *Chlamydia trachomatis* among more than 300.000 men and women aged 16-29 in the Netherlands and did not show a reduction in positivity rates (odds ratio 0.96, 95%CI 0.83-1.10, p-value=0.52) nor estimated population prevalence (3% in the control group vs 2.6% in the intervention group). An Australian cluster randomized controlled trial assessed the effect of yearly screening for *Chlamydia trachomatis* in about 4000 men and women aged 16-29 and did not show a significant reduction in the prevalence of this infection (adjusted relative difference 0.9 (95% CI 0.5 to 1.6; p=0.67).

### Added value of this study

We describe the results of the first randomized controlled trial to compare screening for *Neisseria gonorrhoeae* and *Chlamydia trachomatis* versus non-screening among men who have sex with men taking HIV pre-exposure prophylaxis. In the primary analysis, we found that non-screening was associated with an overall higher incidence of NG/CT infections (IRR 1.318, 95%CI 1.068-1.627), but this difference was driven by non LGV-CT infections alone (IRR=1.435, 95%CI 1.098-1.875) as no difference in NG infections was found (IRR 1.212, 95%CI 0.940–1.564). Given that asymptomatic participants in the non-screening arm were not aware of a positive NG/CT result and thus not treated, two consecutive NG/CT diagnosis in this arm might represent the same, untreated infection. Therefore, we performed a sensitivity analysis, controlling for this ‘untreated-infections-bias’ in the non-screening arm. In this sensitivity analysis, we found no difference in terms of NG and/or CT incidence between both arms. Screening and subsequent treatment for NG/CT was associated with a 21 to 45% increase in antimicrobial consumption.

### Implications of all the available evidence

Our study does not provide strong support in favor of NG/CT screening in MSM taking HIV-PrEP. However, more RCTs are needed to assess the benefits and harms of NG/CT screening in this population.

## Introduction

International guidelines stipulate that screening programs should only be introduced once they have met a set of criteria: the benefits should outweigh the harms, screening should be cost-effective and there should be evidence from high-quality randomized controlled trials (RCT) that screening is effective in reducing mortality or morbidity.^1^ No such RCT has ever been conducted to evaluate the efficacy of screening for *Neisseria gonorrhoeae* (NG) or *Chlamydia trachomatis* (CT) in men who have sex with men (MSM).^2^ Two large cluster RCTs have been conducted to evaluate the effect of screening for CT in general populations.^3,4^ Both found no significant impact of screening on the prevalence of CT. No RCTs have been conducted to evaluate the efficacy of screening for NG.^5^

Ecological analyses have found that countries where MSM are more intensively screened for NG/CT do not have a lower incidence and prevalence of asymptomatic or symptomatic NG/CT cases.^6,7^ One study that used self-reported data from two surveys in 2010 and 2017 of over 100,000 MSM from 46 European countries found that the intensity of NG/CT screening increased over time, but the intensity of screening was positively associated with the number of symptomatic NG/CT cases.^6^ The authors concluded that intensive screening may abrogate the development of an immune response to these infections which paradoxically increases the risk of subsequent re-infection. In the case of CT, there is experimental data from animal models and an observational clinical study to support this ‘arrested immunity’ hypothesis.^8–10^ Although contested, there is also epidemiological evidence that intensive screening of CT in the general population has resulted in lower immunity to CT and subsequent increased incidence.^8^ A number of authors have argued for more frequent NG/CT screening in MSM.^11^ They have largely based this call on modelling studies, some of whom have found that two- to three-monthly screening reduces incidence, and the finding that more frequent screening detects more infections which, if treated, will reduce the population prevalence.^11,12^ Partly as a response to these arguments and evidence of increasing incidence of these infections in many countries, numerous guidelines have increased the recommended intensity of screening for NG/CT to 3-monthly, 3-site (anorectum, urethra and pharynx) testing in MSM taking HIV pre-exposure prophylaxis (PrEP).^13^

We have shown that screening MSM for NG/CT results in high levels of macrolide, cephalosporin and tetracycline consumption.^14,15^ For instance, three-site, three-monthly screening results in up to 12 defined daily doses of macrolides per 1000 inhabitants per year (DID).^14^ This high antimicrobial consumption exceeds the approximate thresholds for the induction of antimicrobial resistance (AMR) in *Streptococcus pneumoniae, Mycoplasma genitalium* and *Treponema pallidum* by 5- to 9-fold.^16^ In other studies, we found a positive ecological association between the intensity of screening MSM for NG/CT and reduced gonococcal susceptibilities to cephalosporins.^17^ Increased antimicrobial consumption is of particular concern in MSM on PrEP as gonococcal AMR has frequently emerged in such core-groups heavily exposed to antimicrobials.^18^ Interestingly, we showed that changing NG/CT screening intensity in a PrEP cohort from three-monthly, three-site to one-site, six-monthly reduced the consumption of macrolides from 12.05 to 3.27 DID without any noticeable adverse clinical consequences.^14^

Given the lack of evidence around screening for NG/CT and the risk of inducing AMR, RCTs are urgently needed to assess the benefits and risks of screening for NG/CT in core groups such as MSM on PrEP. In this paper we present the results from the first RCT to compare the effect of screening on the incidence of NG/CT infections in MSM on PrEP. We also assessed the effect of screening on the incidence of symptomatic NG/CT infections, syphilis infections and antibiotic consumption as well as the PrEP users’ perceptions towards STI screening.

## Methods

### Study design

We performed a multicenter, randomized, controlled clinical trial of three-site three-monthly screening for NG/CT versus non-screening among MSM taking HIV-PrEP in Belgium. The study took place in five HIV reference centers in Belgium (Institute of Tropical Medicine (ITM) in Antwerp, Saint-Pierre University Hospital and Erasme University Hospital in Brussels, Ghent University Hospital in Ghent and Liège University Hospital in Liège). A qualitative sub-study was embedded within the trial at ITM to explore PrEP users’ perceptions towards STI screening. This study was approved by the Institutional Review Board of ITM (1360/20) and by the Ethics Committees of the University Hospital of Antwerp (20/27/377), Saint-Pierre University Hospital (20-07-05), Ghent University Hospital (BC-08167), Erasme University Hospital (P2020/321) and Liège University Hospital (2020-240). The study protocol is available in the Appendix p.6.

### Participants

All men followed-up for PrEP in these five centers were approached for study inclusion. Inclusion criteria were 1) being able and willing to provide informed consent, 2) being born as male, 3) being 18 years old or more, 4) having had oral sex and/or anal sex with another man in the last 12 months, 5) being enrolled in a Belgian PrEP center and 6) being willing to comply with the study procedures. Exclusion criteria were 1) being enrolled in another interventional trial, 2) testing positive for HIV at screening and 3) having symptoms of proctitis or urethritis. Participants provided written informed consent.

### Randomization and masking

Subjects who met all inclusion criteria were randomized 1:1 into the non-screening (intervention) or 3×3 screening (control) arms. The randomization list was prepared by an independent statistician using SAS 9.4 (SAS Institute, Cary NC). To ensure (approximate) treatment balance within study sites, the randomization list was blocked by site using variable block sizes. The overview of the randomization list was not shared with the investigators until trial database lock. Study participants, doctors and nurses were not blinded. The study statistician was blinded until approval of the statistical analysis plan.

### Study procedures

As in routine PrEP care, participants were asked to attend 3-monthly visits at the PrEP clinic. The study duration was 12 months, hence five study visits were planned. One baseline visit took place at day 0 and four subsequent visits at months 3, 6, 9 and 12, each within a window of one week earlier and 6 weeks later.

At the baseline visit, after eligibility assessment, informed consent procedure and randomization, socio-demographic characteristics, sexual behavior, STI history in the past 12 months and antibiotic use in the past 6 months were collected. A first-void urine sample, pharyngeal swab and anorectal swab were collected. The pharyngeal swab was collected by the physician, whereas both other samples were self-collected. Samples per participant were pooled and tested for NG and CT by nucleic acid amplification techniques (NAAT). Those who tested positive were recalled for treatment according to current guidelines.^19^ This generally entailed ceftriaxone 500mg or 1g intra-muscularly with or without azithromycin 2g orally for NG and doxycycline 200mg/day orally for seven days for CT and 21 days for LGV. Syphilis and HIV testing was performed on a blood sample.

At the month 3, 6 and 9 visits, symptoms compatible with an STI, STIs diagnosed, antibiotic use and sexual behavior since the last visit were recorded. A first-void urine sample, pharyngeal swab and anorectal swab were collected from all participants. For asymptomatic participants in the 3×3 screening arm, these samples were analyzed and, if positive, participants were recalled for treatment according to current guidelines. In the non-screening arm, results were only provided when symptoms were present. Asymptomatic participants in the non-screening arms were thus not informed of the result of these samples, nor was the physician who performed the study visit. All participants who reported symptoms either during a study visit, or between study visits were tested and treated as per current guidelines.

At the month 12 visit, data were collected as for the previous visits. A first-void urine sample, pharyngeal swab and anorectal swab were collected and analyzed for NG/CT for all participants. If positive, participants from both arms were treated as per current guidelines. HIV and Syphilis testing was performed on blood samples every 3 months.

Study participants were able to attend the PrEP/STI clinic at any point in between the scheduled visits for any health problems. Participants were encouraged to attend the clinic for any symptoms compatible with an STI. Participants who received a partner notification for an STI were tested and treated according to the current guidelines. Test-of-cure visits were performed according to local protocols.

For the qualitative sub-study, social scientists trained in qualitative research, conducted three focus group discussions (FGD), among randomly selected ITM study participants. Each FGD consisted of three to five participants. To maximize variation in perceptions, two in-depth interviews (IDIs) with PrEP users of the clinic who declined participation to the main study were performed. The interviewers obtained a verbal informed consent from each participant prior to the start of the FGDs and IDIs. Audio-recording took place upon agreement. FGDs and IDIs were conducted in Dutch and online via a secured platform, respecting General Data Protection Regulation.

### Laboratory procedures

NG and CT testing was performed at each site’s laboratory. The three samples were pooled per patient and visit according to a validated pooling strategy^20^. Positive samples for CT were sent to the National Reference Center for STIs (ITM) for genotyping. HIV and syphilis testing was performed according to local protocols.

### Outcomes

The primary outcome was the overall incidence of NG/CT infections in each arm. Each participant could contribute one diagnosis of CT and one diagnosis of NG per scheduled or unscheduled visit. Only laboratory-confirmed diagnoses made between scheduled visits, performed inside or outside of the study clinic were included.

Secondary outcomes were ceftriaxone, azithromycin and doxycycline exposure in the two study arms (expressed in daily defined doses (DDD) per 1000 persons years according to WHO methodology^21^), incidence rate of symptomatic NG and CT and incidence rates of syphilis and HIV.

### Sensitivity and stratified analysis

All NG/CT diagnoses were included in the primary outcome. Hence, it was implicitly assumed that every diagnosis was a new infection. It is however possible that an NG/CT infection detected at the 3 to 12 month visit in the non-screening arm was simply a non-resolved infection that was already present at the prior visit. This could spuriously increase the measured incidence in the non-screening arm as the same infection would be counted twice. Therefore, a sensitivity analysis was performed to deal with this ‘untreated-infection bias’. In this analysis, consecutive diagnoses of the same type (e.g. CT at two consecutive visits) in the non-screening arm were counted as one infection unless the prior diagnosis was a symptomatic one (and therefore treated), or if the participants reported having used antibiotics efficacious against the relevant STI between both diagnoses.

In addition, a pre-specified sub-group analysis was performed by stratifying the participants according to STI risk behavior. We hypothesized that the effects of screening for NG/CT could be different in individuals with a lower number of sexual partners given the lower sexual network connectivity in these individuals. For that purpose, participants that consistently reported 4 or less partners in all 5 study visits were categorized as lower-risk and all other participants were categorized as higher risk. Finally, a separate, non-pre-specified analysis was added using gonorrhoea and chlamydia separately as outcomes.

### Qualitative analysis

All FGDs and IDIs were transcribed verbatim and pseudonymized. Data were collected and analyzed iteratively using a thematic analysis approach and Nvivo. We inductively developed an initial coding scheme. Subsequently, we re-read all transcripts with the focus on describing the variation in perceptions towards testing for asymptomatic and symptomatic NG/CT infections and how the emergence of antibiotic resistance influences these perceptions.

### Statistical analysis

For the primary outcome, estimates were based on a negative-binomial regression model with number of diagnoses as dependent variable, study arm and study site as independent variable and log(visit number) as offset. This model also provided an estimate of the log incidence rate ratio (IRR, no screening versus screening), together with 95% confidence interval. Non-inferiority of the ‘no screening’ arm was concluded if the upper limit of the 95% confidence interval was lower than 1.25. The same methodology was applied for the secondary outcomes except for antimicrobial consumption for which a rate ratio was calculated, with number of DDDs as dependent variable. The number needed to screen was calculated using a previously described methodology.^22^

The primary analysis was performed following the per-protocol (PP) approach. Participants who had fewer than 3 visits with NG/CT results or did not follow the randomized intervention were excluded from the PP analysis. Participants were excluded from the intention to treat (ITT) analysis if they did not attend any of the follow-up visits.

Participants in each intervention arm were described with respect to baseline characteristics. The description was done in terms of medians/means and quartiles/standard deviations for continuous characteristics and using counts and percentages for categorical characteristics.

Assuming that 95% of the participants would have data on all four follow-up visits, and 5% would have data on only three visits, the required sample size to obtain 80% power was 912. Assuming an additional 10% drop out rate, the final sample size was estimated to be 1014 participants.

All statistical analysis were performed using R (version 4.2).

The largest safety concern for this study was that the participants in the non-screening arm could experience a higher incidence of symptomatic NG/CT. Rather than reporting each symptomatic episode of NG/CT as an adverse event, an independent data and safety monitoring board (DSMB) evaluated if the non-screening arm had an unacceptably high incidence of symptomatic NG/CT. For this purpose, the DSMB included two independent STI experts (Infectious Disease Physicians/Epidemiologists) and the study statistician to evaluate the incidence of symptomatic NG and CT in both arms at two interim time points: once 50% and 100% of all study participants had completed their month 6 visit. It was decided that serious consideration would be given to stopping the study if the incidence of symptomatic NG and CT infections in the non-screening arm was double that of the screening arm.

The trial protocol was registered at clinicaltrials.gov (NCT04269434)

## Results

A total of 2409 individuals were approached for the study between the 21st of September 2020 and the 4^th^ of June 2021, among whom 1014 were randomized (508 in the 3X3 screening arm and 506 in the non-screening arm). A total of 38 participants did not attend any follow-up visit and were excluded from the analysis. We excluded 275 participants from the per protocol analysis, 206 had out of window visits, 133 had fewer than three visits with NG/CT results and eight participants in the non-screening arm did not follow the randomized intervention.

Almost all participants identified themselves as cis-male (1011/1014, 99.7%; Table 1) and three individuals identified as trans-woman (3/1014, 0.3%). The median age at baseline was 39 years (IQR 33-47). Participants had a median of four sex partners (IQR 2-8) in the preceding three months. Around one third of the participants (365/1014, 36.0%) reported the use of any antibiotic in the previous six months, 175 (17.3%) reported use of macrolides, 144 (14.2%) use of cephalosporins, 110 (10.8%) use of penicillins and 111 (10.9%) use of tetracyclines. The number of sex partners and unprotected sex partners remained stable across all study visits in both arms (Appendix p.1) The baseline characteristics as well as number of sex partners were well-balanced between the two arms (Table 1).

**Figure 1.**
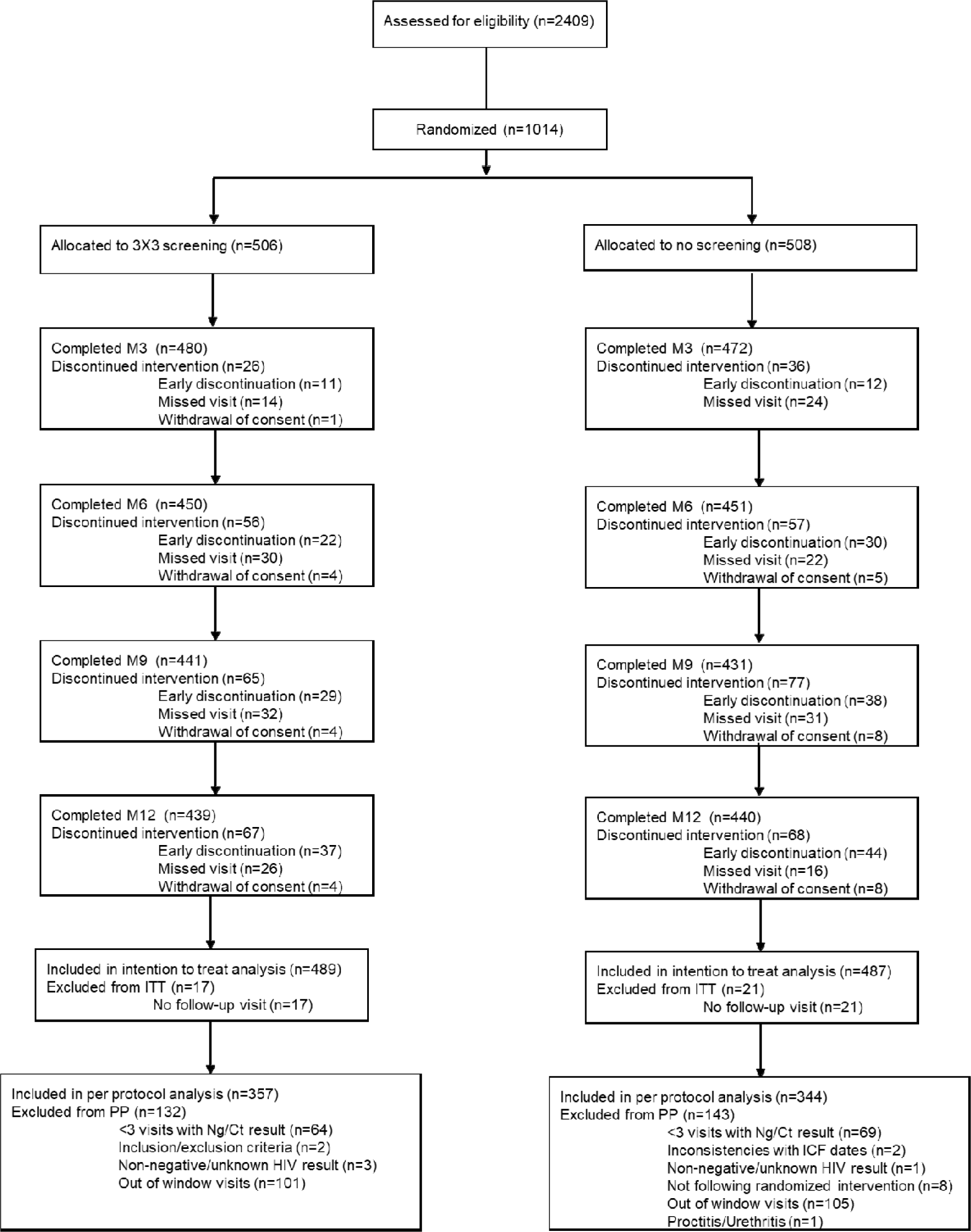
Trial profile

**Table 1.** Baseline characteristics in both arms.

|  | **3 x 3 Screening**<br>**(N=506)**<br>**n (%) / Median (IQR)** | **Non-screening**<br>**(N=508)**<br>**n (%) / Median (IQR)** | **Total population**<br>**(N=1014)**<br>**n (%) / Median**<br>**(IQR)** |
| --- | --- | --- | --- |
| **Age** | 39 (33 - 47) | 39 (32.5 - 48) | 39 (33 - 47) |
| **Sex: Male** | 506 (100%) | 505 (99.4%) | 1011 (99.7%) |
| **Sex: Transwoman** | 0 (0%) | 3 (0.6%) | 3 (0.3%) |
| **Number of sex partners (past 3 months)** | 4 (2 - 8) | 4 (2 - 8) | 4 (2 - 8) |
| **Number of unprotected sex partners (past 3 months)** | 2 (1 - 5) | 2 (1 - 5) | 2 (1 - 5) |
| **Any antibiotic (past 6 months)** | 192 (37.9%) | 173 (34.1%) | 365 (36.0%) |
| **Cephalosporins** | 67 (13.2%) | 77 (15.2%) | 144 (14.2%) |
| **Macrolides** | 81 (16.0%) | 94 (18.5%) | 175 (17.3%) |
| **Penicillins** | 63 (12.5%) | 47 (9.3%) | 110 (10.8%) |
| **Quinolones** | 11 (2.2%) | 5 (1.0%) | 16 (1.6%) |
| **Tetracyclines** | 57 (11.3%) | 54 (10.6%) | 111 (10.9%) |

In the primary analysis, the incidence of NG/CT was 0.205 cases/100 person-days (95%CI 0.171-0.246) in the non-screening arm and 0.155 (95%CI 0.128-0.186) in the 3X3 screening arm (Table 2). The incidence rate (IR) was significantly higher in the non-screening arm compared with the 3X3 screening arm (IR ratio (IRR) 1.318, 95%CI 1.068-1.627, p-value=0.01) and the upper-limit of the 95% confidence interval included the non-inferiority cut-off of 1.25. The incidence rate of symptomatic NG/CT was not significantly higher in the in the non-screening arm (IRR 1.373, 95%CI 0.963-1.956, p-value=0.08; Table 2). Participants in the non-screening arm consumed significantly less azithromycin (RR 0.788, 95%CI 0.719-0.863, p-value<0.01), ceftriaxone (RR 0.561, 95%CI 0.426-0.739, p-value <0.01) and doxycycline (RR 0.55, 95%CI 0.515-0.588, p-value<0.01; Table 3) compared with the 3X3 screening arm. The incidence of syphilis was not significantly higher in the non-screening arm compared with the 3X3 screening arm (IRR 1.471, 95%CI 0.943-2.299, p-value=0.09). There were 14 incident cases of lymphogranuloma venereum (LGV) in the screening arm and 10 cases in the non-screening arm, among which respectively 7 and 3 were symptomatic.

**Table 2.**
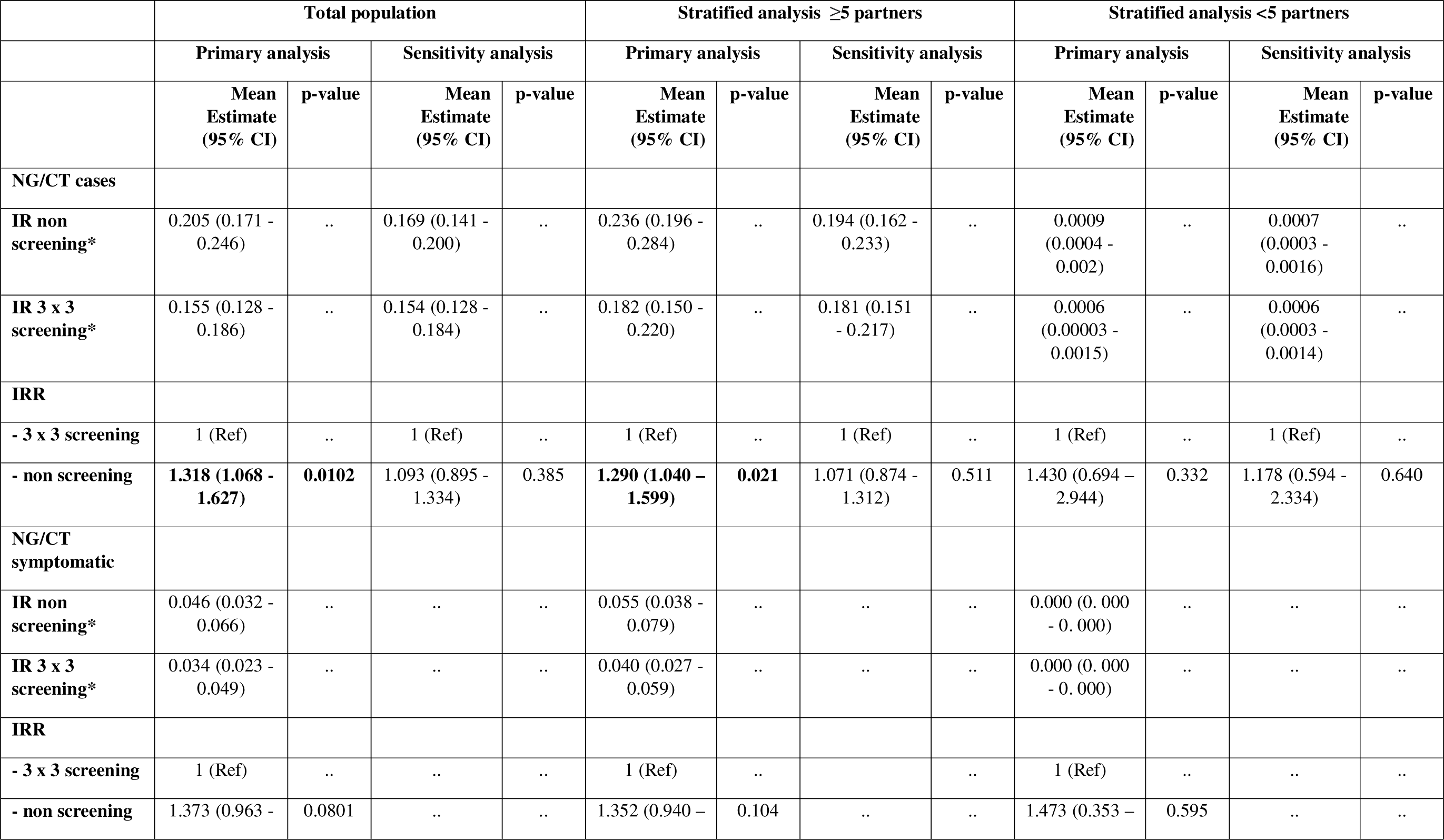
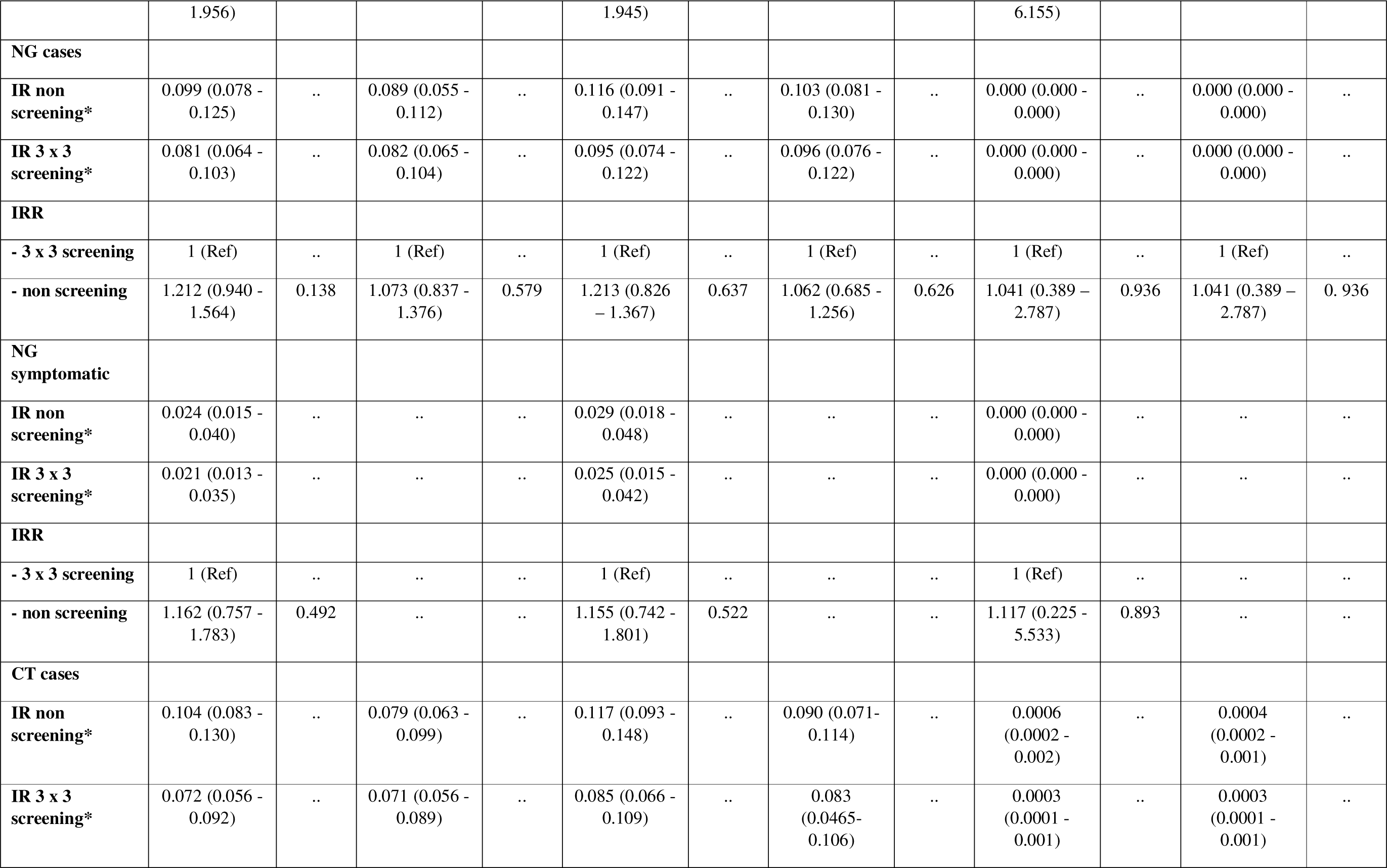
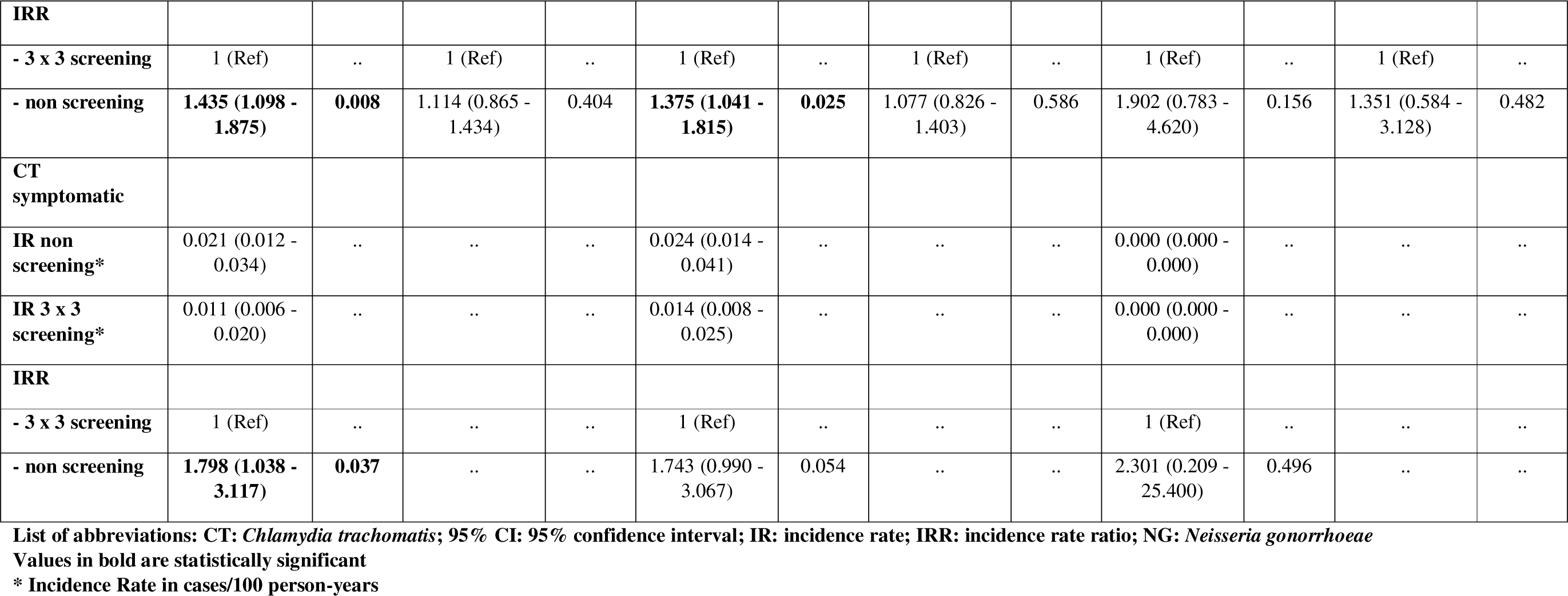
Incidence rate and rate ratio of NG/CT and symptomatic NG/CT (per protocol analysis)

**Table 3.** Rate ratio of antibiotic consumption (per protocol analysis)

|  | Total population |  | Stratified analysis ≥ 5 partners |  | Stratified analysis <5 partners |  |
| --- | --- | --- | --- | --- | --- | --- |
|  | Primary analysis |  |  |  | Primary analysis |  |
|  | Mean Estimate (95% CI) | p-value | Mean Estimate (95% CI) | p-value | Mean Estimate (95% CI) | p-value |
| **Antibiotic consumption** |  |  |  |  |  |  |
| **Azithromycin** |  |  |  |  |  |  |
| non screening* | 0.0046 (0.0043 - 0.0050) | .. | 0.512 (0.367 - 0.713) | .. | 0.139 (0.051 - 0.381) | .. |
| 3 x 3 screening* | 0.0059 (0.0075 - 0.0063) | .. | 0.691 (0.505 - 0.945) | .. | 0.257 (0.096 - 0.689) | .. |
| **RR** |  |  |  |  |  |  |
| - 3 x 3 screening | 1 (Ref) | .. | 1 (Ref) | .. | 1 (Ref) | .. |
| - non screening | **0.788 (0.719 - 0.863)** | **<0.0001** | 0.741 (0.493 - 1.112) | 0.148 | 0.543 (0.124 - 2.208) | 0.393 |
| **Ceftriaxone** |  |  |  |  |  |  |
| non screening* | 0.0004 (0.0004 - 0.0006) | .. | 0.053 (0.041 - 0.068) | .. | 0.015 (0.006 - 0.038) | .. |
| 3 x 3 screening* | 0.0008 (0.0007 - 0.0009) | .. | 0.099 (0.081 - 0.121) | .. | 0.017 (0.007 - 0.038) | .. |
| **RR** |  |  |  |  |  |  |
| - 3 x 3 screening | 1 (Ref) | .. | 1 (Ref) | .. | 1 (Ref) | .. |
| - non screening | **0.561 (0.426 - 0.739)** | **<0.0001** | **0.540 (0.398 – 0.733)** | **<0.0001** | 0.913 (0.312 – 2.677) | 0.869 |
| **Doxycycline** |  |  |  |  |  |  |
| non screening* | 0.0044 (0.0041 - 0.0048) | .. | 0.595 (0.374 - 0.948) | .. | 0.141 (0.031 - 0.644) | .. |
| 3 x 3 screening* | 0.0081 (0.0075 - 0.0086) | .. | 1.028 (0.636 – 1.661) | .. | 0.381 (0.075 – 1.924) | .. |

| RR |  |  |  |  |  |  |
| --- | --- | --- | --- | --- | --- | --- |
| - 3 x 3 screening | 1 (Ref) | .. | 1 (Ref) | .. | 1 (Ref) | .. |
| - non screening | **0.55 (0.515 - 0.0.588)** | **<0.0001** | 0.579 (0.319 - 1.052) | 0.073 | 0.369 (0.034 - 3.991) | 0.412 |

List of abbreviations: 95%CI: 95% confidence interval; RR: rate ratio
Values in bold are significant
\* rate in DDD/100 person-days

In the PP sensitivity analysis accounting for the untreated-infection bias, the incidence rate of NG/CT was 0.169 cases/100 person-days (95% CI 0.141-0.200) in the non-screening arm and 0.154 (95% CI 0.128-0.184) in the 3X3 screening arm. The incidence rate was not significantly different between the two arms (IRR 1.093, 95%CI 0.895-1.334, p-value=0.385) but the 95%CI included the non-inferiority cut-off of 1.25, indicating we cannot conclude non-inferiority of non-screening compared with 3X3 screening.

Results were similar between the PP and ITT analysis, except for the incidence of syphilis that was significantly higher in the non-screening arm compared to the 3X3 screening arm in the ITT analysis (IRR 1.608, 95%CI 1.070–2.418, p-value=0.0224; Appendix p.2).

Differences in NG/CT incidence were driven by differences in CT incidence. There was no difference in NG incidence in the PP analysis (IRR 1.212, 95%CI 0.940–1.5644, p-value=0.138; Table 2) or in symptomatic NG incidence (IRR 1.162, 95%CI 0.757-1.783, p-value=0.579; Table 2). The incidence of CT and symptomatic CT was, however, higher in the non-screening arm (IRR 1.435, 95%CI 1.098-1.875, p-value=0.008; and IRR 1.798, 95%CI 1.038-3.117, p-value=0.037, respectively). There was no difference in CT incidence in the sensitivity analysis (IRR 1.114, 95%CI 0.865-1.434, p-value=0.404). Based on these results, the estimated number needed to screen for symptomatic and asymptomatic CT infections was 27 and 12, respectively (Appendix p.3).

A total of 231 participants reported less than five sex partners at all study visits and where thus considered as lower-risk participants and the remaining 783 participants were considered as higher-risk participants. Higher-risk participants had a significantly higher incidence of NG/CT in the non-screening arm compared with the 3X3 screening arm, in the primary analysis (IRR 1.290, 95%CI 1.040-1.599, p-value=0.021) but this difference was not statistically significant in the sensitivity analysis, when accounting for the untreated-infection bias (IRR 1.071, 95%CI 0.874-1.31, p-value=0.511). The incidences of CT cases and symptomatic CT cases were higher in the non-screening arm in higher risk participants (IRR 1.375, 95%CI 1.041-1.815, p-value=0.025 and IRR 1.743, 95%CI 0.990-3.067, p-value=0.05, respectively). The IRRs in lower-risk participants were not significantly different.

Symptomatic participants typically presented with mild symptoms and no participant reported severe outcomes or adverse events (Appendix p.4). The number of unscheduled visits and visits for partner notification can be found in Appendix p.5.

Participants of the qualitative sub-study reported mixed reactions towards non-screening for asymptomatic NG/CT. The fact that these STIs are mostly asymptomatic and self-limiting, without causing serious complications or harm to the individual, were mentioned as arguments against screening.

> “Why would you try to detect something if you have no symptoms? And that is actually not very dangerous either? Even if you pass it on.” (FGD 3, ID 32)

The main reported disadvantage of non-screening was the possibility of ongoing transmission to sexual partners. For some participants, not testing and treating was accompanied with feelings of guilt, risk, and irresponsibility. Some participants suggested adjusting the testing strategy according to the number of sexual contacts a person has, and whether or not condoms are used.

> “Assuming that a condom is almost never used because there is PrEP. And that there are about five to six or so changing contacts per month. With that in mind, I feel safer being fully tested all the time. If I had a steady partner, and if someone were to come once a month, I would think: okay, let me get tested once every six months.” (FGD 2, ID 26)

The qualitative data showed that perceptions towards AMR varied. Some participants were concerned about the emergence of AMR and/or stated they preferred to avoid using antibiotics when possible. Others reported a lack of knowledge on the subject.

> “I compare it to a scale and I find it difficult to see where that carries the most weight: is the weight in the sense of antibiotic resistance, or is the weight in the sense of I’m walking with an asymptomatic gonorrhoea infection that I could spread to many others. I, personally, find that a difficult balancing act.” (FGD 2, ID 26)

Lastly, not all participants were familiar with the natural course of NG/CT infections and the mechanisms of AMR. As knowledge increased during the sessions, participants’ attitudes sometimes shifted towards non-screening for asymptomatic NG/CT.

## Discussion

This RCT didn’t show that non-screening for NG/CT in MSM on PrEP is non-inferior to 3-site 3-monthly screening with respect to NG/CT incidence. The overall incidence of NG/CT was significantly higher in the non-screening arm compared to the screening arm in the primary analysis. However, in the sensitivity analysis, controlling for the untreated-infections bias, we could not show a statistically significant difference in the incidence of NG/CT between both arms. Differences in NG/CT incidence were driven by a higher incidence of non-LGV CT in the non-screening arm, as the incidence of LGV and NG did not differ. The incidence of symptomatic CT was also higher in the non-screening arm. Participants in the screening arm consumed considerably more antimicrobials compared with the non-screening arm. Among higher-risk participants, the incidence of NG/CT, CT and symptomatic CT were higher as well. These results provide the first RCT-based evidence of the benefits and harms of screening for NG/CT in MSM on PrEP.

Our finding that screening was associated with a lower incidence of CT but not NG is commensurate with the presumed longer duration of infection for CT and possible higher proportion of CT infections that are asymptomatic in MSM.^23,24^ For instance, a systematic review found that chlamydia had a longer duration of infection than gonorrhoea in both the oropharynx and anorectum in MSM.^23^ Hence, periodic screening for NG/CT might detect more CT infections as NG infections might have cleared spontaneously between screening timepoints. While the findings of our study do not provide strong support to continue screening for NG in MSM in PrEP cohorts, they do provide some evidence to support screening for CT. However, as in other studies, we found that the proportion of CT infections that were symptomatic was low (18.4%, Appendix p. 4) and, importantly, we did not find a difference in LGV incidence between study arms.^24^ Nonetheless, it is possible that screening may exert its effect at an individual- and/or population-level. For this reason, it is critical to evaluate the benefits and harms of screening for NG/CT at both levels. Such insights were important for the participants of the qualitative sub-study, as both the risks of ongoing STI transmission and of AMR development were cited as arguments for or against screening.

We have previously established that intense screening for NG/CT is a key driver of high antibiotic consumption in PrEP users.^14,15^ In a similar vein, reducing the intensity of screening for NG/CT in PrEP users has been shown to result in a large reduction in macrolide consumption.^14^ However, screening and subsequent treatment for CT may be less likely to induce AMR than screening for NG. This is because treatment guidelines recommend the less-resistogenic doxycycline for CT therapy compared to NG therapy where ceftriaxone with or without azithromycin (both WHO ‘reserve’ antimicrobials) are advised.^25,26^ We calculated that 12 men would need to be screened at three sites every three months for a year to prevent one asymptomatic CT infection and 27 to prevent one symptomatic CT infection. This would require 2.25 courses of doxycycline therapy for each symptomatic CT infection prevented.

In our study, higher-risk participants had a higher incidence of asymptomatic NG/CT infections. Previous studies have similarly found that the majority of STIs in PrEP cohorts were diagnosed in a small subgroup with a high rate of partner turnover.^27^ In such individuals, the high number of partners results in a dense sexual network which generates a high equilibrium prevalence for STIs such as NG and CT.^28^ Intensive screening for these STIs in this group may reduce this prevalence but would place evolutionary pressures on these STIs to acquire mutations that would enable them to regain their equilibrium prevalence. This could be via evading the diagnostic tests used (as has occurred with CT^8^), or via the emergence of AMR as has transpired on multiple occasions with NG.^18^ Therefore, although the effect of screening for CT was greatest in those with higher STI risk behavior, screening in this group may confer the greatest risk for the emergence of AMR. Modeling studies have suggested that intensive screening may reduce the prevalence of NG/CT to such an extent that the consumption of antibiotics may be reduced in this group.^29^ These modeling studies are, however, at odds with the results of observational studies which have found that the screening MSM for NG/CT was not associated with reduced prevalence regardless of how intensive the screening.^5^

We found an increased incidence of syphilis infections in the non-screening arm compared to the 3X3 screening arm in the ITT analysis. This finding could be explained by the higher consumption of doxycycline and ceftriaxone, two antimicrobials effective against *Treponema pallidum*, in the screening arm. Given that the incubation period of primary syphilis is typically 10-90 days and the fact that syphilis infections are frequently asymptomatic in this population, treating NG/CT with either of these antimicrobials could have reduced the incidence of syphilis.

Our study had several limitations. The untreated-infections-bias meant that our primary analysis overestimated the incidence of NG/CT infections in the non-screening group. Controlling for this bias in our sensitivity analysis may, however, have underestimated NG/CT incidence in the non-screening arm. Moreover, given the number of sex partners reported by participants, there might have been contamination between study arms. Another limitation is that the participants and physicians were not blinded. This might have resulted in altered behavior. This RCT took place in different periods of COVID-19 restrictions. It has been shown that PrEP users decreased their number of partners in the periods of COVID-19 restrictions.^30^ We cannot exclude that our results were impacted by changing behaviors and might thus not be representative of periods with no restrictions.

We conducted this RCT to help inform clinical practice but acknowledge that the results are best interpreted in conjunction with the other types of evidence outlined above. The main reason to screen for NG/CT in MSM is to reduce the incidence of symptomatic infections and secondarily to reduce the incidence/prevalence of infections in the population. In our RCT, screening reduced the incidence of CT but not NG. The effect on CT incidence disappeared once we controlled for the untreated-infections bias. We found that screening resulted in a statistically significant lower incidence of symptomatic CT infections but not symptomatic NG infections. Screening was however associated with a 21 to 45% increase in consumption of antimicrobials. We argue that our results do not provide strong evidence in favor of routine screening of MSM in PrEP programs for NG or CT.

## Authors contribution statement

CK, SH, YVH, IDB, EF, DH, A-SS, J-CG, EP, BV, TR and AL contributed to the conceptualization, methodology and funding acquisition. All authors contributed to the investigation. AT performed the formal analysis. All authors contributed to the writing of the manuscript and approved the final version.

## Data sharing

Anonymized individual participant data and a data dictionary defining each field in the dataset can be shared on approval of a written request to the corresponding author and in agreement with ITM data sharing policy.

## Acknowledgments

The authors thank all study participants, caregivers and investigators involved in the study. This study was funded by the Belgian Healthcare Knowledge Center (KCE).

